# Data-driven Targeting of COVID-19 Vaccination Programs: An Analysis of the Evidence on Impact, Implementation, Ethics and Equity

**DOI:** 10.1101/2023.01.12.23284481

**Authors:** Zoë M. McLaren

## Abstract

The data-driven targeting of COVID-19 vaccination programs is a major determinant of the ongoing toll of COVID-19. Targeting of access to, outreach about and incentives for vaccination can reduce total deaths by 20-50 percent relative to a first-come-first-served allocation. This piece performs a systematic review of the modeling literature on the relative benefits of targeting different groups for vaccination and evaluates the broader scholarly evidence – including analyses of real-world challenges around implementation, equity, and other ethical considerations – to guide vaccination targeting strategies. Three-quarters of the modeling studies reviewed concluded that the most effective way to save lives, reduce hospitalizations and mitigate the ongoing toll of COVID-19 is to target vaccination program resources to high-risk people directly rather than reducing transmission by targeting low-risk people. There is compelling evidence that defining vulnerability based on a combination of age, occupation, underlying medical conditions and geographic location is more effective than targeting based on age alone. Incorporating measures of economic vulnerability into the prioritization scheme not only reduces mortality but also improves equity. The data-driven targeting of COVID-19 vaccination program resources benefits everyone by efficiently mitigating the worst effects of the pandemic until the threat of COVID-19 has passed.

## Introduction

The data-driven targeting of COVID-19 vaccination programs has been a key strategy for mitigating the negative impact of the pandemic so far and is a major determinant of the ongoing toll of COVID-19. Targeted access to, outreach about and incentives for vaccination help maximize the impact of vaccination programs. They eliminate barriers to vaccination and increase uptake among those with the highest infection fatality rate and/or the greatest risk of transmission to others. The return on investment in targeting is enormous.^1^ Data-driven vaccination targeting generates benefits estimated to be on the order of a 20-50% reduction in deaths relative to a first-come-first-served or random allocation.^2-4^

This piece performs a systematic review of the modeling literature on the relative benefits of prioritizing different groups for COVID-19 vaccination and evaluates the broader scholarly evidence on targeting – including analyses of real-world challenges around implementation, equity, and other ethical considerations. Initial and booster vaccination rates among targeted groups are key metrics for measuring the success of ongoing vaccination programs and should guide the allocation of program resources that eliminate barriers to vaccination.

### Two Overarching Strategies

There are two overarching strategies to protect the vulnerable in order to reduce hospitalizations and deaths: vaccinate those who are most vulnerable to severe illness (direct protection) or vaccinate those who are most likely to spread the virus (indirect protection). The benefits of the two strategies can be directly compared in modeling studies but each strategy faces different implementation challenges.

Estimating the impact of direct protection from vaccination is straightforward. A vaccine dose administered to a high-risk recipient who is, for example, 90 times more likely to die than someone in the general population will therefore be 90 times more effective at reducing deaths, on average, than if it were administered to anyone else.^5^ This logic is analogous to calculations of the number needed to vaccinate to prevent one death.

Indirect protection is harder to estimate since there is little data on who contributes most to transmission of the virus. However, on average, an unvaccinated low-risk person likely has more social contacts and contributes more to transmission than an unvaccinated high-risk person, all else equal. The combined effect on severe illness (direct plus indirect) would therefore be lower than the direct effect, but likely not by enough to eliminate the steep gradient in risk of severe illness.

This means a vaccination program would have to more than double the pace of vaccinations to achieve an equivalent impact as targeting the vulnerable who are more than twice as likely to be hospitalized or die. That level of acceleration is unlikely to be possible given other bottlenecks in the system due to a limited supply of implementation resources.

### Systematic review of the modeling literature

A systematic review of the modeling literature was performed to quantify the relative benefits of different COVID-19 vaccine targeting strategies. The review included English language studies that used a mathematical model to examine the impact of COVID-19 vaccination on mortality, morbidity, hospitalization or cases. Studies that did not include real-world data were excluded. A database search of the National Institutes of Health iSearch COVID-19 portfolio identified published manuscripts and pre-prints posted between March 15, 2020 and Dec 15, 2021 that performed SARS-CoV-2 vaccination modeling and included any of the following terms: mathematical, computation, simulation, SEIR, prioritization or optimization. The search produced 1491 papers of which 136 were duplicates. Following a review of the title and abstract of the remaining 1355 studies, 74 were deemed relevant. Of these, 32 were pre-prints in December 2021 but only 17 remain preprints as of December 2022.

The vast majority of these 74 studies concluded that minimizing deaths was a higher priority than minimizing cases, so I focus on those results here. Fifty-five of the 74 relevant studies concluded that when vaccine supply is limited, the most effective way to save lives and return to normal is to vaccinate high-risk people directly, not to reduce transmission by vaccinating low-risk people. (See Appendix A for the full list of 35 of these 55 papers that aren’t cited in this section; see Saadi et al. for a more in-depth narrative review of the modeling evidence.)^6^ Most studies chose to define COVID-19 vulnerability based on age, which is one of the strongest and most well-known predictors of risk, however age is just one of several criteria that should be considered for targeting. For example, certain underlying medical conditions are associated with a higher risk of hospitalization and/or death from COVID-19.^7^ When using the risk of severe COVID-19 illness as the sole criterion for targeting, prioritizing access and uptake among the highest-risk groups to reduce hospitalizations and deaths is optimal. The age-based gradient in severity of illness is so steep that targeting the oldest not only maximizes lives saved but also life-years saved.^8^

The rationale for targeting based on age – the steep gradient in COVID-19 risk – also justifies including additional factors in the prioritization matrix to increase program impact. Two studies noted that there are large benefits to using more than just age in the targeting strategy.^3,9^ Chapman et al. found that targeting by two or more risk factors simultaneously averted up to 40% more disability-adjusted life years (DALYs).^3^

The literature identified several additional factors that could be used for vaccine targeting because they are associated with greater vulnerability to severe illness or higher transmission rates. Ten papers found that prioritizing by both age and occupation (i.e. essential worker status), which takes workplace-related risk differentials into consideration, improves outcomes relative to age-only prioritization.^10-19^ Four studies found that it is optimal to switch to a focus on vaccinating the youngest/lowest-risk once most of the oldest/highest-risk people have been vaccinated.^15,20-22^ Once the majority of the vulnerable are protected, there is a relatively larger benefit to slowing transmission by vaccinating high-contact groups. Five studies found that along with targeting older people, targeting those living in large households, especially ones with elderly members, and those who have more social contacts can help slow transmission to protect the vulnerable.^16,18,23-25^ One study found that, if feasible, prioritizing the elderly who did not have antibodies from a prior COVID-19 infection would be beneficial since prior infection is associated with a reduced risk of severe COVID-19 illness.^26^

By contrast, 18 of the 74 studies found that targeting vaccination of those most likely to spread COVID-19 – including younger age cohorts and/or essential workers – to reduce transmission was a more effective strategy than targeting those most vulnerable to severe illness. Of these, 17 prioritized high contact^27-29^ or high transmission groups including the young,^30-34^ the socially connected,^35,36^ adults in large households and multi-generational homes,^37,38^ migrant workers^39^ or places with high population density^40^ or case growth.^41-43^ One found that targeting based on the presence of SARS-CoV-2 antibodies would be optimal. ^44^

One of the 74 studies found that vaccine targeting mattered little. ^45^

The evidence in favor of targeting high contact or high transmission groups is weaker relative to that targeting high-risk groups since several of the studies made assumptions about vaccine efficacy and/or age-contact patterns that undervalue targeting the elderly and bias the results against the strategy of directly vaccinating the vulnerable. Matrajt et al. assumed a vaccine efficacy of only 60% against severe illness and Mulberry et al. assumed a vaccine efficacy between 60-90% against infection.^29,32^ Several studies use pre-pandemic age-contact patterns that include more age-mixing than during the pandemic.^7^ If the pandemic caused the elderly to be less likely to interact with the young – who are taking more risks since they’re lower risk – then vaccinating the young does relatively less to protect the vulnerable.

Vaccination programs that include frontline essential workers as a prioritization group alongside high-risk individuals will achieve many of the same benefits as strategies that prioritize the young or other high-contact groups. Essential workers, who tend to have more contacts than those working from home, may disproportionately contribute to transmission rates.

Mulberry et al. included estimates of morbidity due to long COVID-19, which places more weight on vaccinating younger people to prevent infections since they would accumulate long COVID-19 morbidity through their lifetime.^29^ Further research is needed to incorporate a consideration of the disability burden into targeting strategies.

### Additional targeting criteria

It is important to ensure that the broader policy debate isn’t constrained by the somewhat narrow focus of the modeling literature on age and essential worker status as the primary targeting criteria. The evidence demonstrates that using multiple criteria at once can improve the effectiveness of targeting. Studies that aren’t model-based nevertheless demonstrate the benefits of targeting by geographic area, socio-economic factors and/or the feasibility of non-pharmaceutical interventions.

It is beneficial to geographically target vaccination access, outreach and incentives to areas such as zip codes with high rates of hospitalization and mortality either by allocating more resources to already-prioritized groups or by expanding the criteria – lowering age thresholds for eligibility for incentives, for example – in high-priority areas.^46^ Geographic targeting produces a multiplier effect: It provides added protection to the most vulnerable in high-mortality areas because transmission is reduced as a greater proportion of the community is vaccinated.^46^

In the U.S., Black, Latino, and Indigenous populations have borne a disproportionate COVID-19 mortality burden. This is mainly due to risk factors, such as underlying medical conditions, essential worker status and lack of access to high-quality health care, that are highly correlated with race.^47^ Incorporating one or more metrics of social disadvantage into the targeting strategy can increase the impact of vaccination programs. Though the social vulnerability index (SVI) alone is not a strong predictor of COVID-19 deaths, it can be used in conjunction with other targeting criteria to improve outcomes.^48^ Allocating vaccination program resources based on the area deprivation index (ADI), that focuses on neighborhood socioeconomic status, can achieve similar benefits.^49^

In some cases, it may be possible to delay vaccination for those who can employ non-pharmaceutical interventions such as masks and social distancing as a viable, albeit more burdensome, substitute to achieve a comparable reduction in transmission and/or severe illness.^50,51^

Ultimately, a high proportion of the population has one or more COVID-19 risk factors so a vaccination targeting program should apply a weighting scheme or use a composite index to determine the relative allocation of vaccination program resources.^52^ A simple and effective solution is to create risk groups based on a combination of a small set of risk factors and then prioritize the groups based on the relative rate of severe illness in each group.

### Implementation challenges and considerations

When crafting real-world policy, policymakers must consider implementation challenges alongside the modeling evidence. A review of the potential implementation challenges provides three points of further support for a strategy of directly vaccinating the vulnerable relative to vaccinating those most likely to spread the virus. First, it’s easier to identify people with characteristics that put them at risk for severe illness – such as age and underlying medical conditions – than to identify those most likely to drive transmission, which depends mainly on the number of social contacts. Using cell phone records to identify the most socially active, as proposed by Hartnett et al., is likely to be legally infeasible.^27^ Targeting those who participate in high-risk activities for vaccination creates moral hazard since it induces people to increase their risky behavior to try to get access to limited vaccination program resources. Second, people at high risk for severe illness have a greater willingness to get vaccinated than those with low risk. Approaches that aim to vaccinate those who contribute most to transmission may be impeded by higher rates of vaccine hesitancy.^53^ Third, a strategy of vaccinating the vulnerable has a clear and predictable pathway from vaccination to intended effect. By contrast, the successful implementation of a strategy of vaccinating to slow transmission hinges on factors with little data such as: vaccine efficacy against transmission, vaccination take-up rates among low-risk groups, and the degree and type of COVID-19 mitigation measures in the future.

Though many studies modeled vulnerability based on age alone, the impact of COVID-19 vaccination programs is increased by including additional risk factors in the targeting, such as occupation, underlying medical conditions and zip code, to ensure those at greatest risk reach the highest vaccination rates. The maximum number of factors is dictated by what is politically feasible and can be communicated clearly and transparently to the general population. Targeting access to, outreach about and incentives for vaccination based on age, occupation, and geographic location (zip code or equivalent) would lead to improved outcomes relative to targeting based on age alone and is clear enough to be understood by the public. For example, Zimmerman et al. proposed vaccines be prioritized to health care workers, then by mortality risk, then by lottery weighted by the ADI.^54^

Some may argue for simple categories to ease logistics, but these are secondary concerns relative to reducing severe illness and can be overcome by better planning and communication. Machine learning or other approaches with complex algorithms are unlikely to be transparent enough to gain the public’s trust and should therefore be avoided.^55^

The successful implementation of any targeting scheme when there are limited resources to ensure access, engage in outreach or provide incentives requires the cooperation of the general public to wait their turn for access to resources rather than try to cut in line. Vaccine prioritization was politically popular during the early stages of the COVID-19 pandemic and survey data demonstrated broad support for prioritization based on age, essential worker status and income levels.^56,57^ Targeting schemes are more likely to be acceptable to the public if they are predictable, fair, and evidence-based.^58^ The potential risk of legal challenges or other significant delays that jeopardize vaccination program implementation should be taken into consideration. For example, including the area deprivation index (ADI) rather than the social vulnerability index (SVI) in the targeting strategy can achieve similar benefits with a far lower risk that legal challenges – even those that are baseless and/or ultimately unsuccessful – could cause enough delays to make that strategy counter-productive.^49^

### Equity and other ethical considerations

The proposed approach of vaccinating the most vulnerable is aligned with standard ethical considerations of saving lives, saving life years and prioritizing the sickest.^59^ It is also in line with the principles of health equity: prevent harm, prioritize the disadvantaged, and aim for equal treatment given equal risk.^55,60^ COVID-19 has widened existing health inequities because it hits already-vulnerable populations the hardest.^47^ That means the principles of saving the most lives and prioritizing the sickest are closely aligned rather than presenting a tradeoff, as often occurs with heath interventions. For example, allocating vaccination program resources based on social vulnerability or area deprivation can increase the number of lives saved by the vaccination program over and above using age alone. So, incorporating them into the targeting scheme not only *improves efficiency* by increasing the number of lives saved but also improves *equity* by increasing the proportion of total lives saved that come from the most vulnerable populations.^46,61^ The overall effect of geographic targeting includes the impact via the multiplier effect: it improves equity in non-targeted vulnerable groups too because they share households and communities.

The ethical obligation to try to maximize lives saved and prevent harm justifies a targeted approach, even if it reduces the overall uptake of vaccination to some degree because the benefits of targeting to high-risk populations are likely to far outweigh the slightly lower uptake rate among low-risk populations. Aiming to raise the overall rate of vaccination uptake alone can be counter-productive, especially given resource constraints, if it leaves vulnerable populations at risk for a longer period because low-risk populations were vaccinated ahead of them. However, increasing the rate and pace of uptake within each targeted group is beneficial, as is allowing some overlap between timing of targeted groups to ensure the system is always operating at full capacity.

## Discussion

It is clear from the literature, including modeling studies and considerations around implementation, equity, and ethics, that targeting the vulnerable for access to, outreach about and incentives for vaccination is preferred to targeting to slow transmission. Given the high toll of the pandemic, the high efficacy of vaccination and the dramatic differences in the risk of severe illness, the return on investment in targeting vaccination resources is enormous.^1-4^ The benefits of targeting are larger when vaccination program resources are more limited and/or fewer transmission mitigation measures are available.^2,4,50,62^

There is compelling evidence that vulnerability should be defined based on more than just age to attain the maximum benefits of from targeting vaccination program resources. For example, a simple metric where vaccination program resources for access, outreach and incentives are proportionally allocated according to groups with highest mortality rates is both efficient and equitable. Creating target groups based on age, occupation group, underlying conditions and geographic location (zip code or equivalent) is simple enough to be feasible yet complex enough to maximize benefits.

There are three gaps in the vaccine prioritization literature that are likely to tilt the balance somewhat towards vaccinating younger populations. Few studies present evidence from low- and middle-income countries which have age distributions that generally skew younger than high-income countries.^6^ Few studies incorporate high-quality measures of the aggregate toll of morbidity, such as quality-adjusted life years (QALYs), that accumulates among COVID-19 and long Covid cases who aren’t ill enough to be hospitalized. Few studies evaluate the impact of vaccination targeting on policy priorities beyond health status, such as education and economic productivity, where the illness and mortality of young people causes a relatively greater loss.

The targeting of access to, outreach about and incentives for COVID-19 vaccination is a major determinant of the continued trajectory of the pandemic. Investing in strategies that efficiently raise the uptake of both initial and booster vaccine doses among targeted groups will maximize the impact of vaccination programs. Data-driven targeting benefits everyone by efficiently mitigating the worst effects of the pandemic until the threat of COVID-19 has passed.

## Data Availability

All data produced in the present work are contained in the manuscript

## Acknowledgements

Melody Pinamang and Yetunde Oshagbemi provided excellent research assistance. The author has no competing interests or funding sources to report.

## Appendix A

List of 35 additional studies not cited directly in the text that concluded that vaccination programs should vaccinate high-risk people directly.

